# Mobilizing community-driven public health response: increasing access to diagnostic testing for underserved and uninsured individuals in Connecticut through lab-in-a-van partnerships

**DOI:** 10.1101/2024.11.21.24317748

**Authors:** Brittany Choate, Ruhani Sardana, Acsah Mathews, Stephanie Weirsman, Katherine Fajardo, Yasmine Ali, Chen Liu, Pei Hui, Kevin Schofield, Anne L. Wyllie, Angelique W. Levi

## Abstract

**Background:** Partnering with community leaders, we sought to address ongoing diagnostic testing needs in underserved neighborhoods and evaluate whether a saliva-based mobile testing program could help overcome barriers to testing for uninsured and low-income individuals. This is critical as many lack a primary care provider, cannot access reliable health information, or have limited financial resources.

**Methods:** Free saliva-based, SARS-CoV-2 diagnostic testing was offered at 123 local community events in Connecticut, between June 2023 to July 2024. The SalivaDirect extraction-free RT-qPCR protocol was run on a CLIA licensed van operated by Yale Pathology Labs under FDA Emergency Use Authorization. Testing locations were identified and advertised in partnership with the community. Patient perspectives on approachability, convenience, and usefulness of mobile testing were recorded via REDcap.

**Results:** Approximately 100 local contacts informed the mobile testing model. Overall, 1,428 individuals participated, with 838 completing a testing experience survey. Of these, 54% identified as Black, Indigenous, People of Color; 59% reported annual household income less than $25,000; 31% were uninsured. Test results were reported in an average 3.1 hours, 48 positive samples were identified. Test takers agreed it was easy to access the van (74%) and felt comfortable (75%); 29% received their first COVID-19 test at the van; 48% were unaware of alternate testing; 44% reported difficulty accessing health care; and 49% identified transportation as a challenge.

**Conclusions:** This study demonstrated the positive impact mobile testing could have for overcoming barriers to accessing healthcare, and its potential to serve as a framework for managing and responding to future public health needs.

## INTRODUCTION

While there is increasing awareness about the health disparities vulnerable communities face,^1^ challenges in addressing them remain. This was exemplified during the COVID-19 pandemic. At 4% of the world’s population, the United States (US) accounted for 25% of the world’s COVID-19 cases.^2^ This catastrophic event exposed fracture lines in the construct of the public healthcare system - specifically, a lack of accessibility to affordable and accurate diagnosis and clinical management.^3^ Disparities emerged among socioeconomically disadvantaged groups, attributed to the lack of health insurance, access to transportation, and individual awareness of the disease severity.^4^ This highlighted the need for effective solutions beyond conventional methods - improved testing, surveillance and monitoring, data transparency, and targeting of public health interventions. As such, we explored a mobile, saliva-based testing program for its potential to overcome testing barriers for typically underserved, hard-to-reach communities with quality diagnostic care. By offering testing at local community sites, we aimed to provide easily accessible, non-invasive, reliable diagnostic testing to uninsured and low-income individuals to allow them to make informed health decisions. In addition, this study served as a model for evaluating community uptake, willingness to test, and overall impressions of mobile testing services to gauge if and how such programs could be incorporated into future public health response efforts.

## METHODS

### Community-driven program design

Community leaders and organizations were engaged throughout the project. Iterative engagement via emails, phone calls, surveys, and in-depth conversations with key stakeholders was used to refine and expand outreach strategies, identify events or services to pair testing events with, and better understand barriers and evolving community health needs. Trusted community contacts were leveraged to build trust with target populations, perform market testing events, and increase participation in mobile testing events. We specifically sought input on testing locations and engagement strategies from community partners (e.g., health departments, community resource distribution centers, food banks, religious institutions, neighborhood associations, etc). In collaboration with community partners, an online scheduling platform was developed to organize testing event requests, and a mobile testing deployment calendar was built to track upcoming events, confirm staffing, and summarize patient testing rates.

### Specimen collection and on-site testing

With support from NIH RADx,^5^ we outfitted a cargo van to provide high-complexity clinical diagnostic testing. Free SARS-CoV-2 testing was offered at community events using the US FDA Emergency Use Authorization SalivaDirect extraction-free RT-qPCR protocol^6^ on a CLIA-licensed van operated under the Yale University School of Medicine’s Department of Pathology with lab personnel from Yale Pathology Labs. Following an IRB-approved protocol [IRB Protocol# 2000034551], individuals were electronically consented by technicians on-site at the mobile van location. After obtaining consent, participants provided a self-collected saliva sample. Samples were tested on the van following the SalivaDirect protocol.^7^ A report with test results was generated for each participant from an electronic health record system and sent to patient-provided emails in a HIPAA compliant manner; should a participant not have access to email, a printed report was made available.

### Survey design and health awareness

After sample provision, participants completed an IRB-approved survey; responses were tracked and securely stored using REDCap electronic data capture tools hosted at Yale University.^8,9^ Survey questions included demographics (e.g. age, income, education level, race) and questions on mobile testing experience (e.g. ease of access, comfort, reason for choosing the mobile testing site); see Supplemental File 1 for complete survey information. Survey responses were exported from REDCap as a .csv file and then imported and analyzed in Excel. Incentives ($10 gift cards) were offered at select testing events to encourage participation in testing and account for the time required to complete the participant survey. During the hours the van was operational at testing sites, a community health worker or navigator was present as a resource for health-related questions. A health worker followed up individuals with positive test results, providing them with information about “at-home” care and precautions, and aiding in accessing local healthcare resources.

## RESULTS

Between June 2023 and July 2024, 123 community testing events were hosted across nine zip code areas in six Connecticut cities - West Haven (56%, 69/123), New Haven (22%, 27/123), New London (15%, 19/123), Shelton (4%, 5/123), Astonia (2%, 2/123), and Bridgeport (1%, 1/123) (Figure 1, Supplemental File 2). Engagement strategy as well as testing event identification was informed by input from approximately 100 organizations and local leaders representing or actively working with underserved neighborhoods in Connecticut. From these events, 1,428 SARS-CoV-2 PCR tests were administered; 92% (725/791) of which were residents of the nine Connecticut zip code (Supplemental File 2). Gift card incentives increased participation over events where gift cards were not offered (0-48 tests per event with gift cards, mean = 12.9; 0-12 tests per event without gift cards, mean = 1.6).

**Figure 1.**
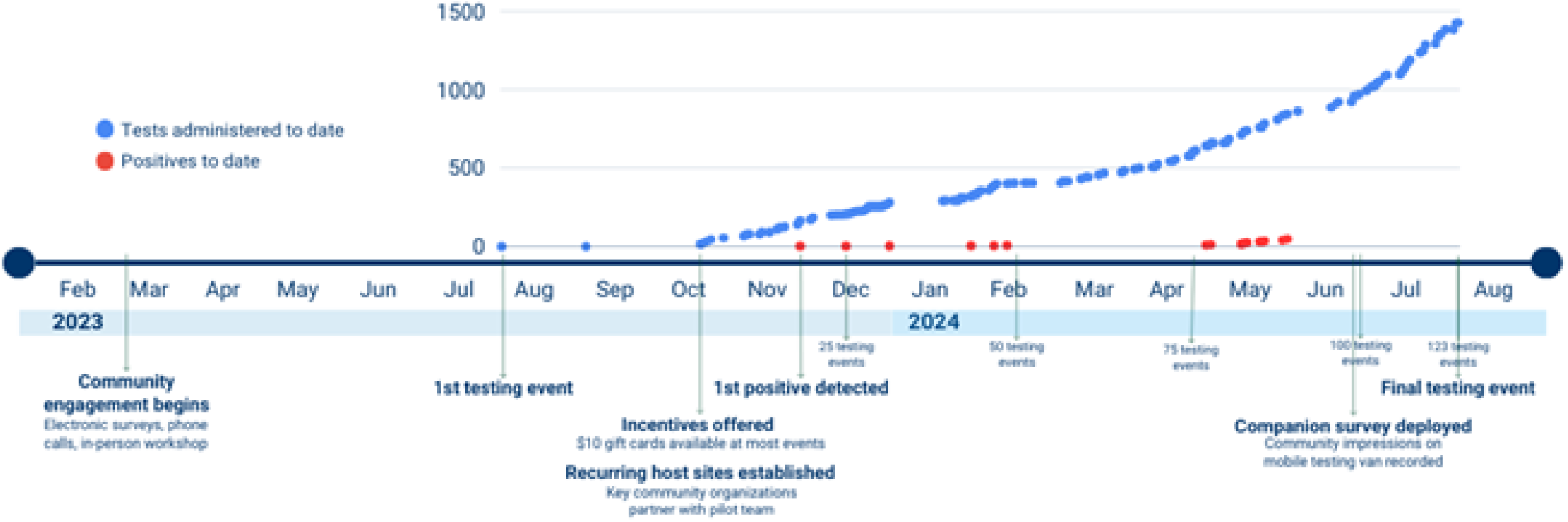
Overview of Connecticut mobile lab project timeline and testing (February 2023 to July 2024). This timeline includes key study milestones (below year line), notably the lead time between the start of community engagement and the first testing event, the gaps between initial testing events, and the increasing frequency as recurring test sites were established. It also shows the increase in tests administered over time (blue) as $10 gift card incentives were introduced and recurring test sites were established and the increase in positive tests over time (red) that the van was able to identify.

### Population demographics

Of the 1,428 patients tested at the mobile van, 838 (59%) responded to the accompanying survey; extent of survey completion varied among test-takers resulting in different statistical denominators as some questions had more respondents than others. Survey respondents ranged from 0-91 years old (mean = 43 years); 54% (302/564) identified as Black, Indigenous, or People of Color; 53% (285/536) identified as male, 41% (218/536) as female, 1% (7/536) as non-binary, and 5% (26/536) preferred not to answer. 59% (285/486) reported annual household income less than $25,000; 91% (442/486) reported less than $50,000. Just over half of the participants were not vaccinated against COVID-19 (55%, 294/540). Of those participants reporting symptoms, 3% (23/641) were experiencing fever, 10% (69/664) cough, 6% (39/664) shortness of breath, and 2% (15/660) loss of taste.

Tests were available irrespective of the insurance status: 31% (251/808) were uninsured; 93% (275/295) of those insured identified “Public (Medicare, Medicaid, Tricare)” as their primary health care plan; only 7% (20/295) said “Private” insurance was their primary plan.

### Test results

Onsite SARS-CoV-2 PCR results were made available to participants in 2-9.6 hours (mean = 3.1 hours). Overall, 48 positive individuals were detected (3%, 48/1428) and received followed ups from a healthcare worker. The majority of tests (80%, 1146/1428) occurred at a single repeat testing site, which averaged 17.1 tests per event (range = 0-48) over 67 separate events - greatly exceeding the average of 5.0 tests/event (range = 0-22) for all other sites.

### Survey output

Survey completion among test-takers varied, resulting in some questions having more respondents than others. The time to complete the survey averaged 7.7 minutes (range = 1.1-39.8 minutes). Survey data showed that: 74% (597/808) of test-takers agreed that it was easy to access the van and get a COVID-19 test; only 5% (42/808) disagreed with this statement and 21% (169/808) were neutral. The majority of test-takers felt comfortable using the van for testing (75%, 608/808); only 5% (37/808) disagreed and 20% (163/808) were neutral. Fewer participants responded when asked why they chose to test at the mobile site, of those who did 58% (272/470) said it was due to “easy access”, 31% (143/470) because they felt comfort and familiarity with the location and/or community representatives involved, 7% (33/470) were referred by either their family, friend or physician; only 5% (21/470) noted the incentive of gift cards as motivating (Table 1; Supplemental File 3). For 29% (167/571) of the cohort, testing at the van was their first COVID-19 test. The questionnaire documented that 48% (255/529) were unaware of alternate COVID-19 testing opportunities; 44% (218/493) reported difficulty in accessing health care; and 49% (241/492) identified transportation as a challenge.

**Table 1.** Responses to survey questions, by demographic group, regarding opinion of the comfort and ease of testing access as well as primary reason for selecting the van for testing (Connecticut, USA; July 2023 to July 2024).**

| Demographic | "Overall, you were comfortable using the van to get a Covid test today." | "Overall, you felt it was easy to access the van and get a COVID test today." |
| --- | --- | --- |
| Age Range, years (n) |  |  |
| 0-17 (50) | Strongly agree | Strongly agree |
| 18-34 (90) | Strongly agree | Strongly agree |
| 35-44 (111) | Strongly agree | Strongly agree |
| 45-54 (77) | Strongly agree | Strongly agree |
| 55-64 (90) | Strongly agree | Strongly agree |
| 65-74 (45) | Strongly agree | Strongly agree |
| 75+ (8) | Somewhat agree | Somewhat agree |
| Gender (n) |  |  |
| Woman (218) | Strongly agree | Strongly agree |
| Man (285) | Strongly agree | Strongly agree |
| Non-binary (7) | Strongly agree / Neutral* | Strongly agree / Neutral* |
| Prefer not to answer (26) | Strongly agree | Strongly agree |
| Race (n) |  |  |
| American Indian or Alaska Native (30) | Somewhat agree | Neutral |
| Black or African American (210) | Strongly agree | Strongly agree |
| Asian (14) | Strongly agree | Strongly agree |
| Native Hawaiian or Other Pacific Islander (1) | Strongly agree | Strongly agree |
| White (262) | Strongly agree | Strongly agree |
| Some other race (47) | Strongly agree | Strongly agree |
| Prefer not to answer (39) | Strongly agree | Strongly agree |
| Annual household income, 2019 before taxes (n) |  |  |
| \$0 - \$14,999 (167) | Strongly agree | Strongly agree |
| \$15,000 - \$19,999 (77) | Strongly agree | Strongly agree |
| \$20,000 - \$24,999 (41) | Somewhat agree / Strongly agree* | Somewhat agree |
| \$25,000 - \$34,999 (71) | Somewhat agree | Somewhat agree |
| \$35,000 - \$49,999 (86) | Strongly agree | Strongly agree |
| \$50,000 + (44) | Strongly agree | Strongly agree |
| Prefer not to answer (88) | Strongly agree | Strongly agree |

\* Equal response (n)
\*\*The extent of survey completion varied among test-takers resulting in rows having different statistical denominators.

| Demographic | "Why did you choose this location for Covid testing?" |
| --- | --- |
| Age Range, years (n) |  |
| 0-17 (37) | Access |
| 18-34 (32) | Comfort/familiarity |
| 35-44 (84) | Access |
| 45-54 (35) | Access |
| 55-64 (33) | Access |
| 65-74 (21) | Access |
| 75+ (6) | Access |
| Gender (n) |  |
| Woman (151) | Access |
| Man (198) | Access |
| Non-binary (5) | Access |
| Prefer not to answer (11) | Access |
| Race (n) |  |
| American Indian or Alaska Native (9) | Access |
| Black or African American (152) | Access |
| Asian (13) | Referral |
| Native Hawaiian or Other Pacific Islander (1) | Access |
| White (197) | Access |
| Some other race (29) | Access |
| Prefer not to answer (18) | Access |
| Annual household income, 2019 before taxes (n) |  |
| \$0 - \$14,999 (123) | Access |
| \$15,000 - \$19,999 (54) | Access |
| \$20,000 - \$24,999 (33) | Access |
| \$25,000 - \$34,999 (57) | Comfort/familiarity |
| \$35,000 - \$49,999 (51) | Comfort/familiarity |
| \$50,000 + (39) | Access |
| Prefer not to answer (53) | Access |

## DISCUSSION

Diagnostic testing has remained a cornerstone in managing the global COVID-19 pandemic since early 2020. However, even several years after the first reported case, testing sites for COVID-19 are not equitably distributed across the United States^10^; often underrepresented minority populations, who arguably have the highest need for testing, bear the brunt of this improper management.^11^ Hurdles are not just confined to the number of testing centers, but also various factors at societal and individual levels, such as lack of access to testing or reliable healthcare information and limited financial resources.^12^ Testing should be accessible and ideally intertwined with people’s everyday routines, livelihoods, and interpersonal relationships. To combat health inequity, we must address unmet needs and remove barriers to accessing health services, such as limited transportation, knowledge, or insurance.

As such, we worked in partnership with the community to curate testing implementation strategies and deploy a community-engaged study that addressed barriers to testing. We sought to explore mobile testing as a model for community-engaged COVID-19 testing by leveraging existing partnerships to accelerate implementation and build trust; iterative engagement and inclusive efforts to understand critical barriers to testing in communities at increased risk for COVID-19; and innovative methods like on-site mobile van testing to support community participation. Beyond COVID-19, it was hoped that this model and resulting strong academic-community relationships could serve as a framework to shape efficient responses to future public health crises.

### Based on the survey conducted as part of this study, participants identified the following as barriers to testing

#### Inadequate resources

Our sites were open to everyone free of charge and regardless of insurance coverage or referral and were free of charge, something not often the case at this stage in the COVID-19 pandemic response. This made a significant difference for our participating population as 91% had an annual household income of less than $50,000 and 31% of our test seekers were uninsured. Of those insured, 93% identified “Public (Medicare, Medicaid, Tricare)” as their primary health care plan. In a study reported by Bevan *et al*.,^13^ both lack of insurance and uncertainty in reimbursement were frequently mentioned as deterrents to seeking care. This barrier may be why, even after testing being available for years, 29% of participants had their first-ever COVID-19 test at the van.

Beyond the ability to pay for a test, 44% reported that just accessing needed health care was a challenge. However, if they could access testing, they reported that fear of the consequences of testing positive introduced other barriers. No clear policies are available by the government on the “best next step” for seeking healthcare if symptom(s) surface or a positive result is returned, the ones provided are widely variable including home isolation, taking leave from work, and reporting to urgent care. These measures can be impractical for under-resourced families, where multi-generational families may live under the same roof or working from home is not an option. Though most of the participants lived alone (56%); 35% (160/458) of the participants lived in a nuclear family, and 9% lived in a multi-generation household or shared the accommodation with other adults. It was concerning to see that 41% did not think they could safely quarantine or isolate themselves without risking their jobs. This calls for better guidelines to support those who test positive, including paid time off from work and quarantine shelter facilities.^14^ Furthermore, Knight *et al*. discussed the reasonable fear of losing government provided shelter in populations experiencing homelessness, which can lead to strong resistance to getting themselves tested.^15^

#### Engagement hurdles

It is not just about the test and the testing, barriers also include uncertainty about eligibility, difficulty interpreting symptoms, and logistical challenges such as transport to and from test sites. During our survey collection, it was surprising that 51% were not aware of alternative testing options in their community, 32% reported testing at the van as their first COVID-19 test, and almost half of the cohort (49%) identified “transportation” as a challenge.

Participants described how unclear messaging from state and national government hindered testing uptake as it was difficult to understand complicated testing eligibility criteria, and self-recognizing symptoms.^13,16^ Bureaucratic or technical barriers to accessing testing also play a role, for example, problems with internet access and/or technological literacy.^17^ The negative association between institutional distrust and healthcare utilization among racial/ethnic minority populations is well-documented and has continued to build during the COVID-19 pandemic.^18^ This broken bridge can be repaired by involving more community healthcare institutions and outreach programs. Government mistreatment of immigrants (e.g., fears of Immigrant and Customs Enforcement) further exacerbates these challenges.^19^

Moreover, every society and ethnic group has social stigma and/or superstitions associated with testing positive for COVID-19.^2,20^ This directly impacts the type and tone of messaging needed to communicate public health information - simply translating materials is not enough, they must be trans-created. Members of the public have consistently reported confusion and uncertainty regarding how to access or where to go for testing, suggesting that public awareness of testing systems is an important factor in testing uptake.^16^ In contrast, our mobile testing model was recognized as convenient and accessible by 79% of the test takers; only 5% disagreed with the van being convenient and accessible for COVID-19 testing demonstrating that our mobile model can effectively offer positively received diagnostic testing traditionally underserved communities.

### Addressing barriers through community-driven public health

#### Establish relationships with trusted community partners

The mobile testing pilot clearly showed the benefits of partnering with established organizations and local leaders for the success of a community testing program. By leveraging existing relationships and on-the-ground knowledge, resources, opportunities and existing complementary programs were able to be more readily identified. This not only expedited data gathering and program development but increased the effectiveness of the testing program and ensured that what was built appropriately addressed existing challenges and gaps in the community’s public health response efforts. Robust community partnerships built by the project team were instrumental in the success of the mobile pilot.

Study partners that made the biggest impact were those that had deep community connections. This was a significant advantage for reaching target demographics but also came with the challenge of working with organizations and individuals that already had existing demands on their time. When engaging with these partners it was critical to anticipate the support they would need and identify and remove barriers. Solutions proven through this study included providing printed advertising materials, setting up onsite signage, assisting with virtual event promotions, and ensuring that the mobile testing team was self-sufficient when onsite - arriving with all the supplies necessary from a power source to test kits. Keeping partnerships as low of a lift as possible meant that key connections continued to engage because the return they saw for their community outweighed the cost of their participation.

#### Embrace iterative engagement

Ongoing, consistent engagement with partners led to a more diverse and effective community testing program. For the pilot, this began with the initial program design where known contacts were asked for input on the program and to recommend additional individuals or organizations that should be involved in the mobile testing program. Broadening the network involved meant that the community cohort was far more diverse, leading to better informed engagement plans and a study design more representative of the target community’s needs, which set up the pilot for greater success. Continuing to engage through emails, virtual meetings, and phone calls between the program team, partners, and host sites improved program response times and the overall testing experience. Checking in and inviting feedback allowed for more dynamic program management, such as updating outreach materials, adding new testing events, or actively responding to patient feedback. Through this, we also learned the importance of maintaining a routine schedule and the impact of offering small incentives for participation, neither of which have been addressed in similar previous studies. Both helped build trust, generate community referrals and, over time, resulted in a significant increase in participation. Furthermore, planning events at repeat locations also increased operational efficiency, allowing for templated event planning and building trust with the population.

#### Build a testing program with target demographics in mind

In line with data reported by others,^13, 21-24^ we observed an overall altruistic take towards testing for COVID-19. Test-taker feedback reported a high agreement on the ease of use and comfort with accessing saliva-based testing at local community events. The design of a mobile testing program must take into consideration its target demographics. Understanding of the challenges and opportunities can be achieved by collaborating with community leaders and organizations who have an established relationship with the neighborhood and are aware of the demands, deficits, and strengths of existing resources. Survey results demonstrate mobile testing as a promising approach for delivering clinical diagnostic testing to the target population and reaching the target demographics. This indicates that mobile programs should be considered as a practical strategy for future testing needs of underserved populations, whether for screening or surveillance efforts or during health emergencies such as outbreak response efforts. Aligning events with existing resources or public health services and deploying local testing events in higher-density areas increased uptake. Testing was most successful at repeat, demographic-aligned locations. For example, 58 testing events were held to coincide with a weekly food distribution event, which accounted for 80% of the tests administered throughout the study and five times the number of tests per event than all other sites.

#### Ensure there is value in what is being offered to the community

Reviewing community needs and listening to feedback from trusted partners throughout the study was critical to its success. In response to the evolving community priorities and assigned “value”, we revised the program design in response. For example, community feedback confirmed that declining interest in SARS-CoV-2 testing and the extensive participant survey were both significant barriers limiting participant enrolment. To address this, we started offering $10 gift card incentives. As a result, we saw a significant and immediate increase in the level of participation.

## PUBLIC HEALTH IMPLICATIONS

Our mobile testing implementation, designed to improve access to COVID-19 testing, is a model others can draw from to rapidly provide health services to communities that are under-resourced. Information collected through our surveys confirms that mobile testing is a feasible, accessible, and flexible way to meet the healthcare needs of the community and is a model shown to be highly accepted by the public. This study also confirmed that our method of engaging target populations through a community-driven approach can lead to high participation levels in a testing experience that was easy to use and comfortable for participants. This indicates that community-driven mobile programs should be considered as a practical strategy for future testing needs of underserved populations. Furthermore, this intervention demonstrates the potential for strong nonprofit––academic–community partnerships to form and rapidly respond to community health needs. By combining resources from local nonprofits and well-equipped medical or academic institutions with the on-the-ground expertise of community-centered organizations, we can work together to jointly address structural and systemic inequities key to realizing health equity.

This community-focused study demonstrated the positive impact a mobile testing program could have in addressing unmet needs and removing barriers to accessing health services, such as limited transportation, knowledge, or insurance. Collaborating with community representatives is essential in implementing effective diagnostic testing programs in underserved populations. The findings of this study suggest that testing should be understood as a social event that is intertwined with people’s everyday routines, livelihoods, and interpersonal relationships; thoughtful program design, listening to the target community, and responding to dynamic needs are critical.

## Data Availability

All data produced in the present study are available upon reasonable request to the authors

---

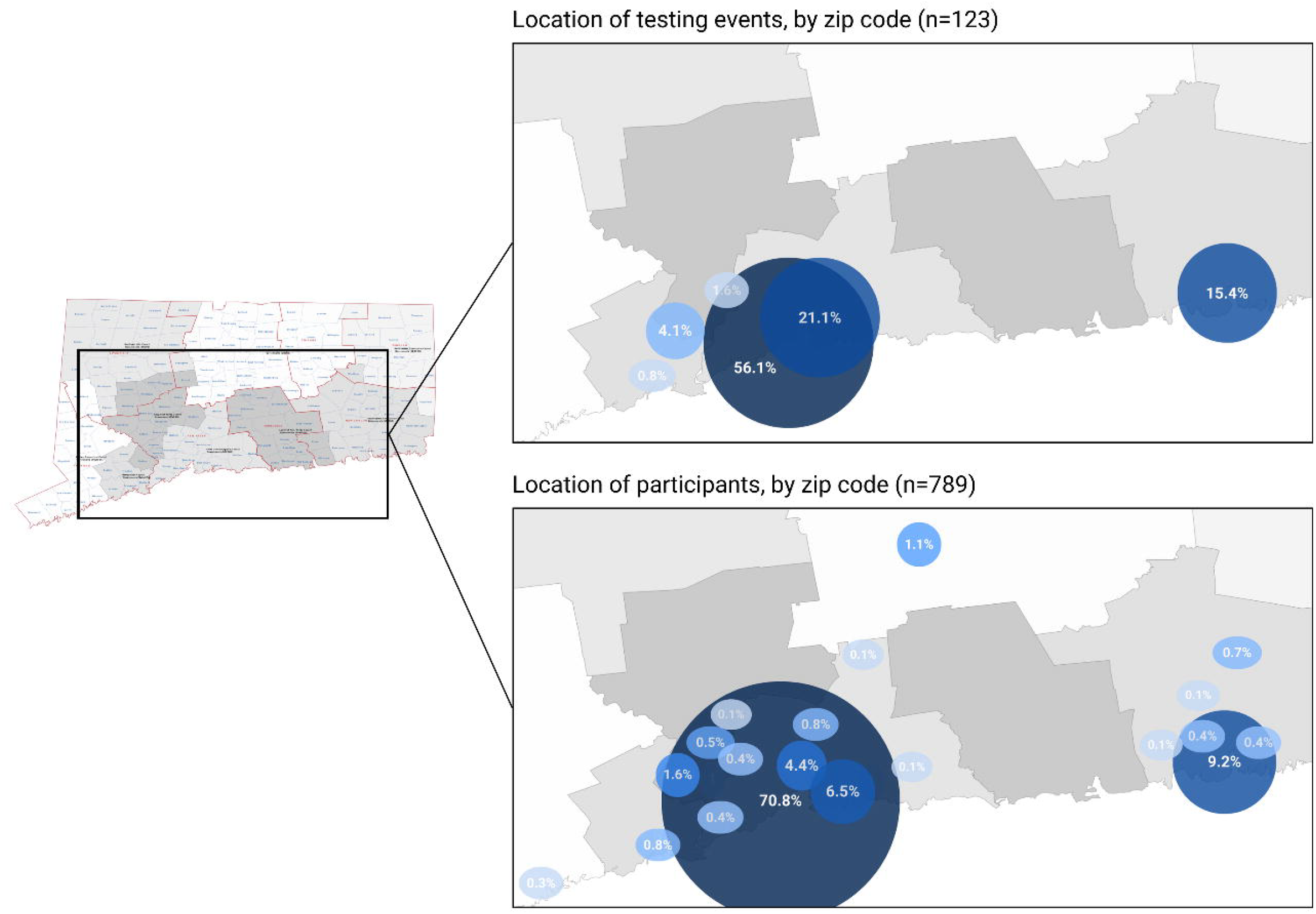

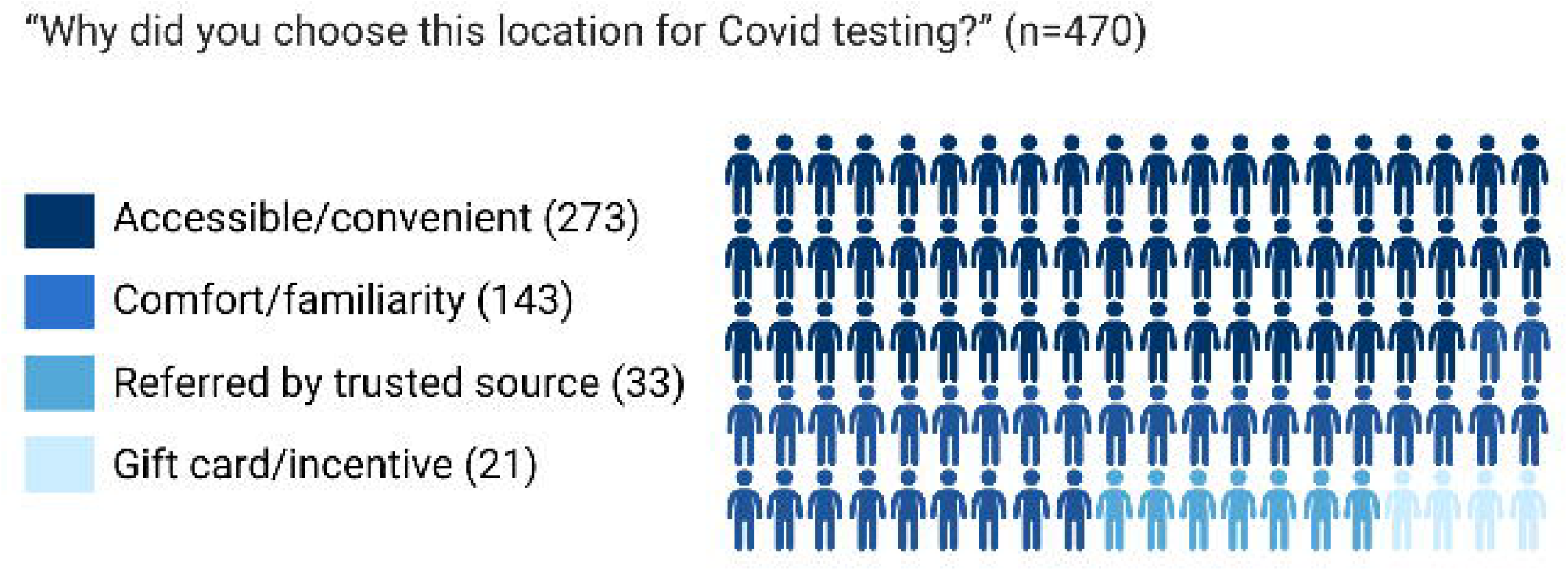

