## Supplemental File 1; Supplemental File 2 for "Mobilizing community-driven public health response: increasing access to diagnostic testing for underserved and uninsured individuals in Connecticut through lab-in-a-van partnerships"

### SUPPLEMENTAL FILES

**Supplemental File 1.** The IRB-approved survey questions asked of Connecticut mobile testing study participants (July 2023 to July 2024) were taken directly from the NIH RADx-UP Common Data Elements (Phase 3) standardized question set (NIH RADx-UP CDEs v1.6 Tier 1 REDCap Codebook). To review the Phase 3 REDCap Codebook and see a complete list of survey questions, visit: https://radx-up.org/wp-content/uploads/2022/12/RADx-UP_1.61_Phase3_Tier1-_Codebook.pdf

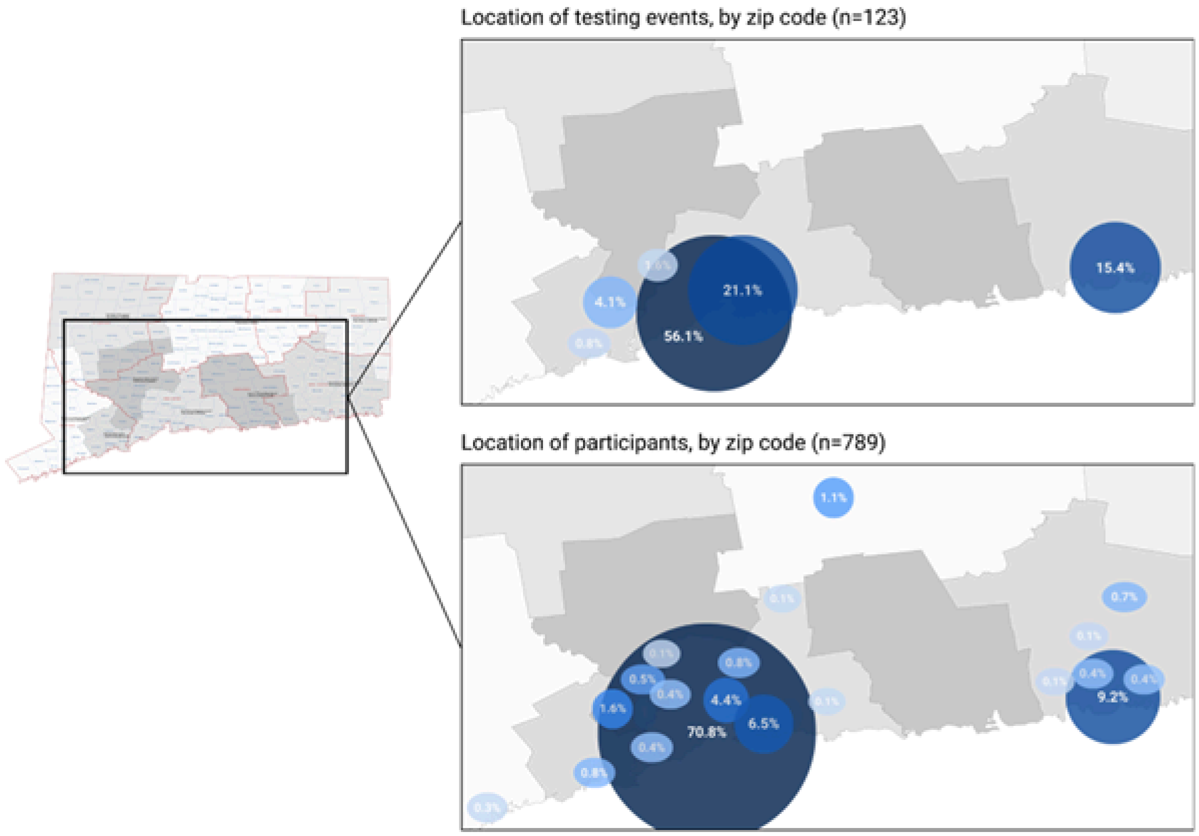


***Supplemental File 2.*** *Location of Connecticut mobile testing events and participants (July 2023 to July 2024). These maps show the significant overlap (92% agreement) between the geographic area, by zip code, of testing events (top) and the zip code of mobile testing participants (bottom). Circle sizes and colors are representative of the relative number of each (darker = larger number); corresponding percentages are inset.*

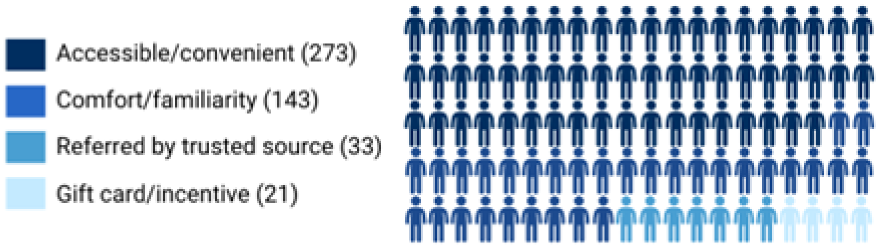


***Supplemental File 3. Responses to the question: "Why did you choose this location for Covid testing?" (n=470).*** *More than half of participants in Connecticut chose to test at the mobile van because it was accessible/convenient, followed by comfort/familiarity. A relatively small group cited referrals as their main reason for testing at the mobile community site. The least identified reason for testing at the can was the gift card/incentive offered.*
